# REVAMP (Reducing Experimentation of Vaping AMong secondary school Students): Protocol for an Intervention Development Study

**DOI:** 10.1101/2025.09.01.25334882

**Authors:** Andrew Radley, Vicky Moynihan, Alison Duncan, Martine Miller, Angela Niven, Linda Bauld, Kurt Ribisl, Fiona Dobbie

## Abstract

**Background:** Electronic cigarettes (‘vapes’) and other nicotine products such as oral pouches are increasingly used by adolescents. Consumption of these products by young people creates the potential for nicotine addiction and future use of tobacco, as well as associated respiratory problems including coughing, wheezing, and lung inflammation. At present, there are very few evidence-based vaping and nicotine prevention interventions targeting young people in the UK secondary school setting. Considering this gap, combined with rising normalisation of vaping in young people, the proposed development study aims to address a growing public health concern by creating a secondary school-based vaping and nicotine prevention intervention.

**Materials and methods:** REVAMP will involve three phases. Phase 1 will include a review of existing literature on a) school-based electronic cigarettes/vaping prevention programmes globally, and b) causal and contextual factors associated with young people’s vaping. Reviews will include grey literature searches alongside synthesis and updates of existing systematic reviews. Phase 2 will involve a qualitative study, conducting semi-structured interviews and focus group discussions with secondary school students (n=16-20, age 11-13), school staff (n=6-8), parents and carers, and other stakeholders (n=20-26). These activities will explore the views and experiences of existing and future school-based approaches to vaping and nicotine consumption. In Phase 3 workshops will be held with students, school staff, parents and carers and wider stakeholders to consolidate findings from Phases 1 and 2 and collectively develop programme theory and intervention activities. Fieldwork for Phases 2-3 will take place in two secondary schools with profiles of high and low socioeconomic deprivation, respectively, within a single Scottish city. Findings will be disseminated through journal articles, conference presentations, local stakeholder events, and social media.

**Discussion:** REVAMP presents a much-needed and timely pathway towards developing an evidence-based, school-based vaping prevention programme for young people in Scotland. This intervention will complement UK-Wide regulatory efforts to reduce vaping and nicotine product access and appeal for young people. Strengths of this development study include engagement of a wide range of stakeholders, co-creation activities with young people, and extensive exploration of the international evidence base.

## Introduction

Electronic cigarettes (commonly referred to as ‘vapes’) are battery-operated devices that heat a liquid containing nicotine, flavours, and other chemicals to produce an aerosol that is inhaled by the user. While vapes have been proven to be an effective smoking cessation and harm reduction tool for adult smokers, the rapid rise in vaping among adolescents is widely recognised as an urgent public health priority (1-4). By 2024, the proportion of young people aged 11-17 in Great Britain who had tried vaping had more than doubled during the past decade and is now estimated at 18% (ASH, 2024). The Scottish Government’s most recent Health and Wellbeing Census (2021-22) found that vaping was a regular health-harming behaviour in the lives of many young people, with 10.1% of 15-year-olds and 4.3% of 13-year-olds reporting vaping at least once a week (4). Significant inequalities exist, with young people in the most deprived areas of Scotland almost twice as likely to regularly vape than those in the most affluent areas. Girls are also more at risk, 13% of 13-year-old girls reported current vaping compared to 6% of boys (3).

Alongside vaping, other forms of nicotine consumption are also gaining popularity among young people. Products such as oral nicotine pouches (sometimes called snus) are increasingly used, with 2024 data suggesting that 3% of Scottish 11-15-year-olds had tried pouches, reflecting the growing popularity of these products elsewhere in Europe (5). Poly-use of multiple nicotine-containing products, including vapes, cigarettes and pouches, is also common among young people who have developed nicotine dependency (6-8).

Vaping and nicotine consumption in young people raises several health concerns, with the long-term effects unclear. One of the primary issues is the potential for nicotine addiction. Evidence suggests that adolescent nicotine consumption can result in abnormal brain development, poor concentration and memory, impulsive behaviour, and increased risk of mental health problems in adulthood (9-11). Additionally, adolescent vaping has been linked to respiratory conditions such as asthma, chronic obstructive pulmonary disease, bronchitis, and cardiovascular effects (12-15).

The use of disposable vapes has contributed to the sharp rise in adolescent vaping and nicotine consumption, with 54% of UK vapers aged 11 to 17 using disposable vapes in 2024 (2). The recent introduction, by the UK and devolved governments, of new laws to ban disposable vapes may reduce the ease with which young people access vapes. However, these measures are unlikely to completely interrupt access, with recent research identifying social sourcing of vaping products from peers, older friends, siblings and other informal purchases identified as among the most common routes through which young people access vaping devices (2, 17). Unintended consequences such as a potential rise in the illicit trade of disposable vapes; uptake of new EC (vape) products coming onto the market to enable industry to bypass restrictions around disposable vape products and reframing refillable vape products as “eco-friendly” (such as refillable vapes, prefilled pod vapes, and rechargeable vape pods); increases in cigarette smoking and use of other nicotine products (such as snus/nicotine pouches) also raise concerns (18, 19).

Schools remain a popular setting for delivery of interventions to prevent risk taking behaviour for several reasons, not least because school attendance is compulsory which means that the majority of children can be reached through school (20). Existing school-based vaping prevention interventions are in various stages of development and evaluation (21). Early evidence suggests that peer-based approaches and strengthening of social and emotional skills may be effective in preventing or delaying vaping experimentation (22, 23). However, at present, there is no evidence-based vaping prevention intervention targeting young people in secondary schools in the UK.

The REVAMP study therefore will conduct the development work required to co-create programme theory with young people to inform a secondary school intervention to prevent vaping and nicotine consumption among adolescents.

## Methods

### Aim and research objectives

The REVAMP study aims to inform the development of an evidence-based, school-based intervention to prevent vaping and nicotine product consumption among adolescents. This aim is addressed through the following objectives:

1. To identify and critically appraise existing school-based vaping prevention interventions reported in the international literature, assessing their relevance and transferability to the UK context.
2. To investigate the messages perceived to be effective in delaying or preventing the uptake of vaping and nicotine products among adolescents.
3. To explore stakeholder perspectives on the optimal delivery of vaping and nicotine prevention messaging within school settings.
4. To engage young people and key stakeholders in the development of programme theory and intervention components to inform a future trauma-informed, school-based prevention programme.

### Study design

In response to the lack of evidence-based vaping prevention interventions in the literature (21-23), REVAMP will be developed using an established intervention development framework - Six Steps in Quality Intervention Development (6SQuID) (24). This study will follow the first four steps of 6SQUID that seek to: 1) define and understand the problem and its causes; 2) clarify which causal and contextual factors are malleable and have the greatest scope for change; 3) identify mechanisms of change, and; 4) identify how to deliver the change mechanism (24). Steps 5-6 focus on testing, refining and evaluating the intervention and are beyond the scope of this development study.

A multi-modal research design comprised of three phases, summarised in Figure 1, will be conducted.

**Figure 1.**
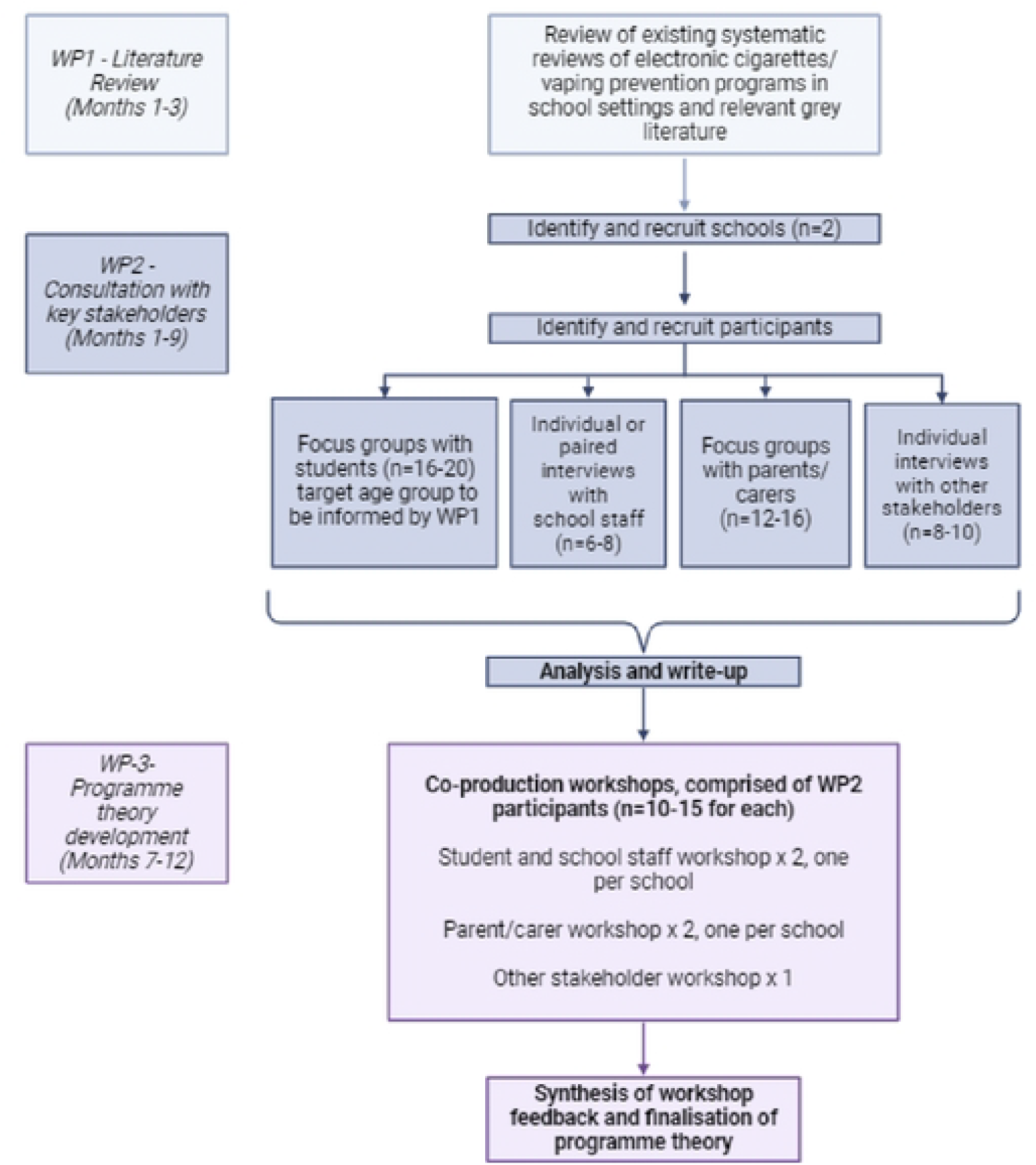
REVAMP study workflow

An advisory board including public health professionals, education professionals, third sector colleagues, policymakers, youth workers, and parents will be convened and will meet biannually to guide the study. The study is sponsored by ACCORD (Academic and Clinical Central Office for Research and Development) and research ethics permission granted by Edinburgh Medical School Research Ethics Committee (EMREC) (REC Number: 24-EMREC-074).

### Phase 1: Literature Review

Phase 1 will address research objective 1 by reviewing existing literature on vaping prevention interventions for young people, including both peer-reviewed and grey literature. Two reviews will be conducted. The first will seek to identify and characterise school-based vaping prevention interventions internationally. The second will examine the causal and contextual factors associated with adolescent vaping.

### Review of School-Based Vaping Prevention Interventions

We will synthesise findings from two recent systematic reviews (22, 23) and conduct an updated rapid review to capture relevant studies published after June 2023. Searches will be conducted in PubMed, Web of Science, Scopus, APA PsycINFO, and the Cochrane Database of Systematic Reviews, using search terms relating to vaping, prevention, young people, and schools. Inclusion will be limited to English-language publications post-May 2023.

Grey literature will be identified through: (1) structured Google searches, (2) websites of relevant organisations known to the research team, and (3) recommendations from the project advisory board and via stakeholder interviews. Data will be extracted on intervention characteristics (e.g., target population, setting, theoretical basis, content, delivery, evaluation methods and findings) and summarised using narrative synthesis (25).

### Review of Causal and Contextual Factors

A rapid review of reviews will be conducted to identify factors influencing adolescent vaping. Searches in Cochrane and PubMed will target English-language literature reviews from January 2020 onward, focusing on adolescents (aged 10–18) in high-income countries. Grey literature will be identified using similar methods. Identified factors will be thematically synthesised and mapped using a socioecological framework (26), categorising influences as intrapersonal, interpersonal, community, or societal.

### Phase 2: Qualitative stakeholder consultation

Phase 2 will involve qualitative interviews and focus groups with key stakeholders including young people, school staff, parents and carers, and professionals working within and across the fields of public health, policy, health care, voluntary sector and education). Phase 2 will address research objectives 2-3. The planned timetable for project delivery is set out in Figure 2 below

**Figure 2:**
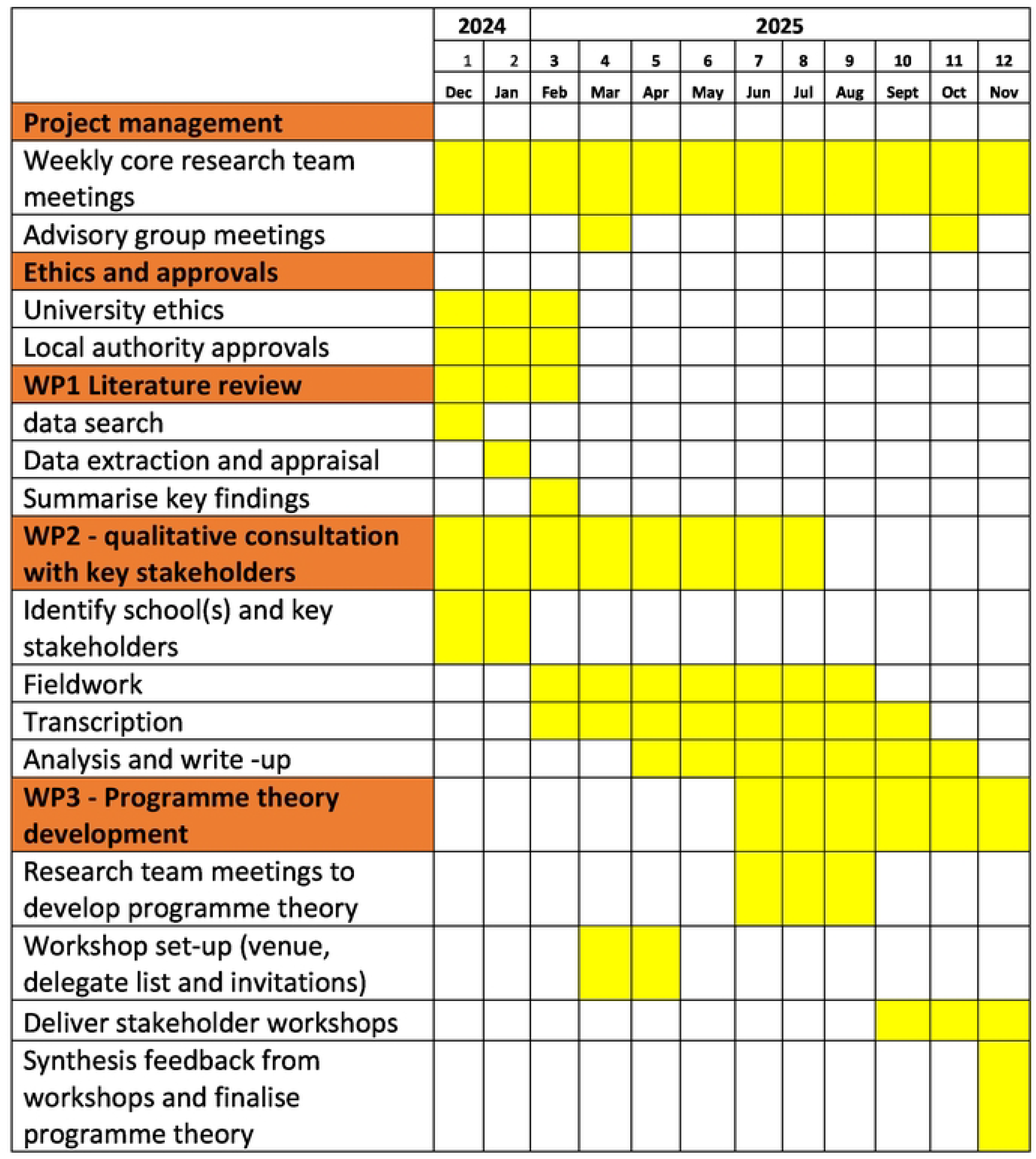
Planned Timetable for Project Delivery

Data collection will take place using a mixture of semi-structured interviews and focus groups depending on the participant group (Table 1). Secondary school students will be invited to take part in small focus group discussions conducted face-to-face in school. Prior to participating, students will provide written assent. Interviews will be guided by a topic guide. This will be developed based on the learning from Phase 1, but will broadly explore young people’s knowledge of vaping and nicotine products, health beliefs and risk perceptions, perceived influences on vaping behaviour, and preferences for content and delivery of a vaping and nicotine prevention intervention. In response to the recent UK ban on disposable vapes (16), focus groups will also explore potential unintended consequences of this ban such as increased need for nicotine withdrawal support among young people, and increased uptake of other nicotine products such as alternative EC (vape) products, tobacco cigarettes or oral nicotine pouches.

**Table 1.**
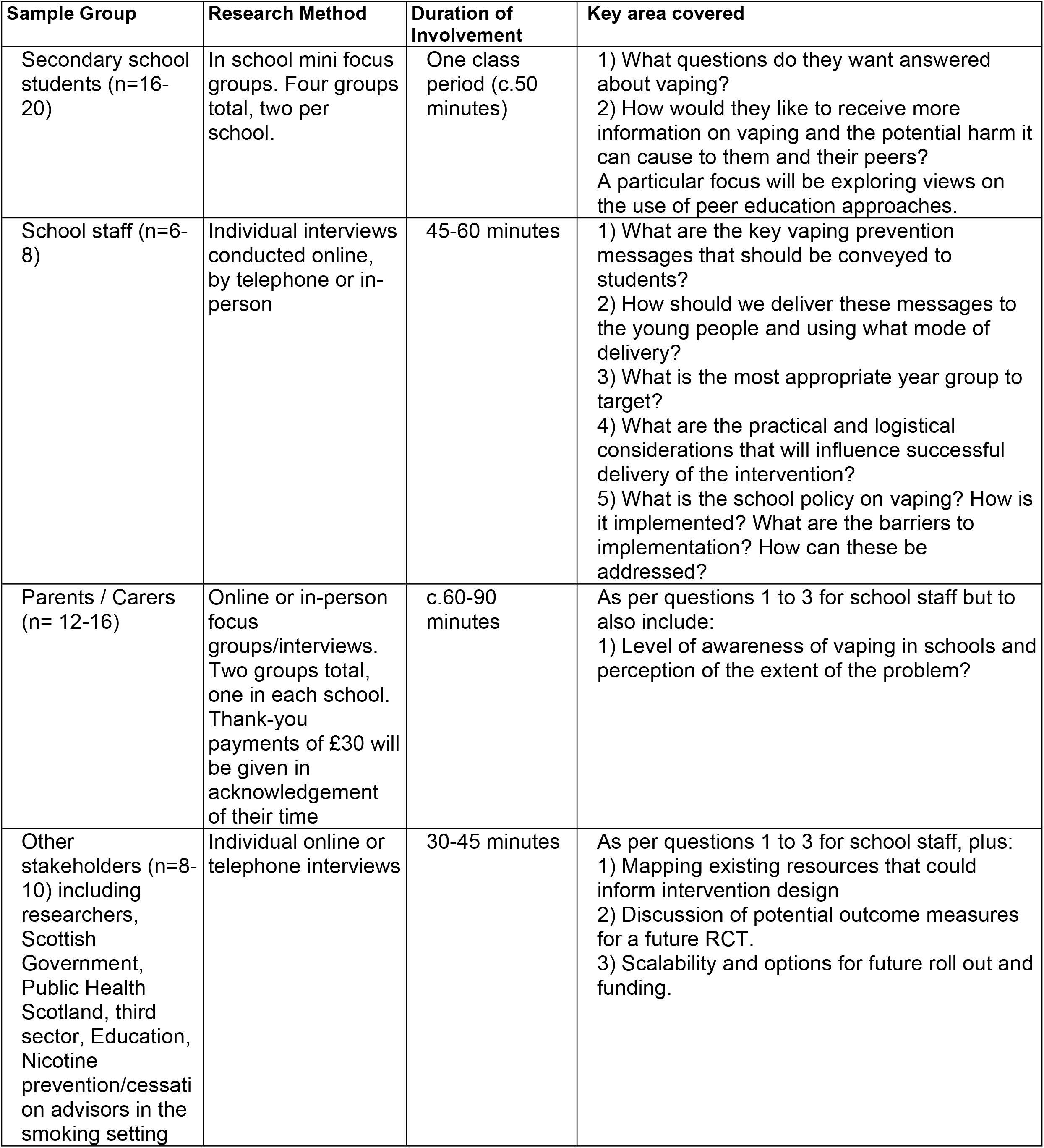
Phase 2 Qualitative Data Collection

| Sample Group | Research Method | Duration of Involvement | Key area covered |
| --- | --- | --- | --- |
| Secondary school students (n=16-20) | In school mini focus groups. Four groups total, two per school. | One class period (c.50 minutes) | 1) What questions do they want answered about vaping?<br>2) How would they like to receive more information on vaping and the potential harm it can cause to them and their peers?<br>A particular focus will be exploring views on the use of peer education approaches. |
| School staff (n=6-8) | Individual interviews conducted online, by telephone or in-person | 45-60 minutes | 1) What are the key vaping prevention messages that should be conveyed to students?<br>2) How should we deliver these messages to the young people and using what mode of delivery?<br>3) What is the most appropriate year group to target?<br>4) What are the practical and logistical considerations that will influence successful delivery of the intervention?<br>5) What is the school policy on vaping? How is it implemented? What are the barriers to implementation? How can these be addressed? |
| Parents / Carers (n= 12-16) | Online or in-person focus groups/interviews. Two groups total, one in each school. Thank-you payments of £30 will be given in acknowledgement of their time | c.60-90 minutes | As per questions 1 to 3 for school staff but to also include:<br>1) Level of awareness of vaping in schools and perception of the extent of the problem? |
| Other stakeholders (n=8-10) including researchers, Scottish Government, Public Health Scotland, third sector, Education, Nicotine prevention/cessation advisors in the smoking setting | Individual online or telephone interviews | 30-45 minutes | As per questions 1 to 3 for school staff, plus:<br>1) Mapping existing resources that could inform intervention design<br>2) Discussion of potential outcome measures for a future RCT.<br>3) Scalability and options for future roll out and funding. |

Individual, semi-structured, interviews will be conducted with school staff to explore: perceptions of what influences young people’s vaping behaviour, current school responses; what should be included in a future intervention; how a future intervention should be delivered; and exploration of potential barriers or facilitators to intervention implementation. School staff will be offered a choice of face-to-face interviews at their place of work or online or telephone interviews. Other stakeholders including healthcare professionals, policymakers, education, and third sector workers will also be invited to take part in semi-structured interviews. These interviews will take place remotely and will follow a similar topic guide, with the addition of questions around scale-up and rollout of future interventions.

Focus group discussions will be conducted with parents and carers following a similar topic guide as for school staff, with additional focus areas around parental awareness of vaping and existing policies/school responses to the issue of students vaping within school grounds, and potential for involving parents and carers in a whole-school approach to vaping. Discussions are expected to last approximately 60-90 minutes, hosted online or face-to-face according to participant preferences. Participants in the parent and carer sample will be given a £30 shopping voucher to thank them for their time and reimburse any expenses invoked in attending the focus group discussion.

### Phase 3 - Programme Theory Development

Phase 3 will focus on programme theory development (research objective 4). Following the initial steps of the 6SQuID framework, data collected from phases 1 and 2 will be reviewed and the causal factors prioritised for intervention development. We will then consider the mechanisms of action (i.e., the processes through which the intervention produces change), identify the specific components of the intervention, and determine the modes of delivery.

Phase 3 will involve working with young people, school staff, parents and carers and wider stakeholders to share findings, explore construction of a theory of change and discuss the potential for unintended consequences, both positive and negative (27). We will convene five parallel co-production workshops, two for school students and staff, two for parents and carers, and one for non-school-based stakeholders (e.g., public health professionals, academics, policymakers). Informed by evidence gathered from phases 1 and 2, the purpose of the workshops is to refine programme theory and intervention delivery mechanisms. Workshops with school staff and students will take place face-to-face at participating schools, during the school day. Parent, carer and stakeholder workshops will take place online or face-to-face at a school or local community venue, according to participant preferences with an option for day or evening delivery.

### Setting

The setting will be two Scottish secondary schools, located within the same local authority area and in an urban setting. While vaping is widespread among Scottish adolescents, those residing in more socioeconomically disadvantaged areas are almost twice as likely to vape (4).Reflecting this, we will recruit one school in an area of high deprivation and another in an area of low deprivation using the Scottish Index of Multiple Deprivation (SIMD) to identify schools (28).

### Sample and recruitment

Target sample groups are school students (11-13 years, n=16-20), school staff (n=6-8), parents/carers (n=12-16) and stakeholders working in public health, education, local and national government (n=8-10). Two schools will be recruited via the research team’s existing networks. Special schools, residential schools and private schools will be excluded. Students in the target year group and their parents or carers will receive an information sheet and a parental opt-out form in advance of fieldwork. Participants will be drawn from students who have not been opted out of the study, based on students volunteering to participate and input from their teachers to select a more diverse and representative sample. School staff and parents and carers will receive information sheets in advance of the fieldwork and will contact the school lead to express their interest in participating.

Stakeholders will be identified using the research team’s existing professional networks, with input from the project advisory board. Snowball sampling will be used to enhance recruitment.

### Analysis and dissemination

All interviews and focus group discussions will be audio-recorded using an encrypted recording device and transcribed verbatim. We will use a thematic approach to analyse the data, facilitated by NVivo 14 software. First, we will read the transcripts to identify the key topics and issues which emerge from the data. Next, a draft analytical framework will be created, piloted, refined and finalised by the project team. Each transcript will then be coded and summarised into key themes using framework matrices, or charts (29). This approach reduces large volumes of data and facilitates a systematic approach between and within case analysis. The use of NVivo 14 ensures that analysis is fully documented, and conclusions can be clearly linked back to the original source data.

Findings will be disseminated via academic and social media routes. We will also seek stakeholder advice on how and where to disseminate our findings beyond peer-reviewed articles, in particular the development of guidance to support young people wishing to find out more about stopping the use of vapes and nicotine products.

## Discussion

The rising normalisation of vaping among young people in recent years is an urgent public health concern. While school-based vaping prevention interventions are in various stages of development and evaluation internationally (21), to our knowledge there are no existing evidence-based interventions developed specifically for the UK or Scottish context. Furthermore, the existing international evidence base is still in its infancy and has been criticised for a lack of robust, theoretically-informed intervention development and rigorous evaluation (21-23). REVAMP will address this gap by developing programme theory to underpin an evidence-based, contextually relevant vaping prevention intervention for the secondary school setting.

The development of REVAMP is timely. The school-based, behavioural change approach used by REVAMP will complement the recent UK regulatory interventions aimed at curbing youth vaping. These include a national ban on disposable vaping products (16) and the Tobacco and Vapes Bill (which will restrict the sale of vaping and tobacco products to people born after 1 January 2009, and introduce enhanced marketing and licencing restrictions for retailers) (30). While these regulatory changes are welcome, they are unlikely to be sufficient to curb youth vaping with many young people accessing vaping products through peers and unofficial sales channels (2, 31). School-based interventions have been shown to be effective at preventing and delaying young people’s experimentation with various other substances (32, 33) and offer an opportunity to reach large numbers of young people, and contributing to the promotion of healthy environments for young people to grow up in (34), thus positioning REVAMP as a much-needed non-regulatory intervention to complement these policy changes.

### Strengths and Limitations

Strengths of this study include a robust research design with literature searches to map existing school-based vaping prevention interventions internationally for adaptation, thus minimising duplication. A further strength, is the wide range of stakeholders represented in the qualitative study. By including the voices of young people at multiple stages of this development study, the outcomes from this early work should translate into more effective interventions (35). Finally, the inclusion of schools from varying degrees of socioeconomic disadvantage within this study will inform the development of a more equitable, trauma-informed vaping prevention intervention.

A key limitation of this study is the small number of participating schools and their shared location within one urban area. The exclusive focus on qualitative work also means that only a small number of young people will participate in development research. The voluntary nature of this participation may introduce sampling bias, with greater interest in participation expected from health-conscious young people who already have negative attitudes towards vaping. To combat this, teachers will be consulted to aid selection of a maximally representative sample, and the study population will be continually monitored and discussed by the research team.

## Conclusion

The use of vape devices has become a pressing issue for young people, parents and families, educators, health workers and policy makers. REVAMP presents a much-needed and timely pathway towards developing an evidence-based, school-based vaping prevention programme for young people in the secondary school setting.

## Data Availability

Data collected during this study will be stored in a pseudonymised format on a University of Edinburgh server. Access to the final trial dataset will be restricted to the Chief Investigator and research team. Anonymised study results will be made available upon reasonable request

## Notes

### Competing Interest Statement

The authors have declared no competing interest.

### Funding Statement

Yes

### Author Declarations

The study is sponsored by ACCORD (Academic and Clinical Central Office for Research and Development) at University of Edinburgh

## References

1. Salari N, Rahimi S, Darvishi N, Abdolmaleki A, Mohammadi M. The global prevalence of E-cigarettes in youth: A comprehensive systematic review and meta-analysis. Public Health Pract (Oxf). 2024;7:100506.

2. Action on Smoking and Health (ASH). Use of vapes (e-cigarettes) among young people in Great Britain. 2024.

3. Inchley J MJ, Brown J, Willis M, Currie D. (2023) MRC/CSO Social and Public Health Sciences Unit, University of Glasgow. Health Behaviour in School-aged Children (HBSC) 2022 Survey in Scotland: National Report.; 2023.

4. Scottish Government. Health and Wellbeing Census 2021-2022. 2022.

5. Brose L, Bunce L, Cheeseman H. Prevalence of Nicotine Pouch Use Among Youth and Adults in Great Britain-Analysis of Cross-Sectional, Nationally Representative Surveys. Nicotine Tob Res. 2025. Epub. 2025 Jan 10.

6. Post A, Gilljam H, Rosendahl I, Bremberg S, Galanti MR. Symptoms of nicotine dependence in a cohort of Swedish youths: a comparison between smokers, smokeless tobacco users and dual tobacco users. Addiction. 2010;105(4):740–6.

7. Gomez Y, Creamer M, Trivers KF, Anic G, Morse AL, Reissig C, Agaku I. Patterns of tobacco use and nicotine dependence among youth, United States, 2017-2018. Prev Med. 2020;141:106284.

8. Azagba S, Shan L, Latham K. Adolescent Dual Use Classification and Its Association With Nicotine Dependence and Quit Intentions. J Adolesc Health. 2019;65(2):195–201.

9. Yuan M, Cross SJ, Loughlin SE, Leslie FM. Nicotine and the adolescent brain. J Physiol. 2015;593(16):3397–412.

10. Goriounova NA, Mansvelder HD. Short- and long-term consequences of nicotine exposure during adolescence for prefrontal cortex neuronal network function. Cold Spring Harb Perspect Med. 2012;2(12):a012120.

11. England LJ, Aagaard K, Bloch M, Conway K, Cosgrove K, Grana R, et al. Developmental toxicity of nicotine: A transdisciplinary synthesis and implications for emerging tobacco products. Neurosci Biobehav Rev. 2017;72:176–89.

12. Kaur G, Pinkston R, McLemore B, Dorsey WC, Batra S. Immunological and toxicological risk assessment of e-cigarettes. Eur Respir Rev. 2018;27(147).

13. Tzortzi A, Kapetanstrataki M, Evangelopoulou V, Beghrakis P. A Systematic Literature Review of E-Cigarette-Related Illness and Injury: Not Just for the Respirologist. Int J Environ Res Public Health. 2020;17(7).

14. Benyo SE, Bruinsma TJ, Drda E, Brady-Olympia J, Hicks SD, Boehmer S, Olympia RP. Risk Factors and Medical Symptoms Associated With Electronic Vapor Product Use Among Adolescents and Young Adults. Clin Pediatr (Phila). 2021;60(6-7):279–89.

15. Gonzalez JE, Cooke WH. Acute effects of electronic cigarettes on arterial pressure and peripheral sympathetic activity in young nonsmokers. Am J Physiol Heart Circ Physiol. 2021;320(1):H248–h55.

16. Scottish Government. Environmental Protection (Single-use Vapes) (Scotland) Regulations 2024: enforcement guidance. 2025.

17. Becker TD, Rice TR. Youth vaping: a review and update on global epidemiology, physical and behavioral health risks, and clinical considerations. Eur J Pediatr. 2022;181(2):453–62.

18. McConnell R, Barrington-Trimis JL, Wang K, Urman R, Hong H, Unger J, et al. Electronic Cigarette Use and Respiratory Symptoms in Adolescents. Am J Respir Crit Care Med. 2017;195(8):1043–9.

19. Watts C, McGill B, Rose S, Yazidjoglou A, Chapman L, Dessaix A, Freeman B. ‘It’ll save your lungs’: early insights into nicotine pouch use and perceptions among young Australians. Health Promot Int. 2024;39(6).

20. White J, Hawkins J, Madden K, Grant A, Er V, Angel L, et al. Public Health Research. Adapting the ASSIST model of informal peer-led intervention delivery to the Talk to FRANK drug prevention programme in UK secondary schools (ASSIST + FRANK): intervention development, refinement and a pilot cluster randomised controlled trial. Public Health Research; 2017. https://www.journalslibrary.nihr.ac.uk/phr/PHR05070.

21. Barnes C, Turon H, McCrabb S, Hodder RK, Yoong SL, Stockings E, et al. Interventions to prevent or cease electronic cigarette use in children and adolescents. Cochrane Database Syst Rev. 2023;11(11):Cd015511.

22. Mylocopos G, Wennberg E, Reiter A, Hébert-Losier A, Filion KB, Windle SB, et al. Interventions for Preventing E-Cigarette Use Among Children and Youth: A Systematic Review. Am J Prev Med. 2024;66(2):351–70.

23. Gardner LA, Rowe AL, Newton NC, Egan L, Hunter E, Devine EK, et al. A Systematic Review and Meta-analysis of School-Based Preventive Interventions Targeting E-Cigarette Use Among Adolescents. Prev Sci. 2024;25(7):1104–21.

24. Wight D, Wimbush E, Jepson R, Doi L. Six steps in quality intervention development (6SQuID). Journal of Epidemiology and Community Health. 2016;70(5):520.

25. Popay J, Roberts H, Sowden A, Petticrew M, Arai L, Rodgers M, et al. Guidance on the conduct of narrative synthesis in systematic reviews: A product from the ESRC Methods Programme 2006.

26. McLeroy KR, Bibeau D, Steckler A, Glanz K. An ecological perspective on health promotion programs. Health Educ Q. 1988;15(4):351–77.

27. De Silva MJ, Breuer E, Lee L, Asher L, Chowdhary N, Lund C, Patel V. Theory of Change: a theory-driven approach to enhance the Medical Research Council’s framework for complex interventions. Trials. 2014;15(1):267.

28. Scottish Government. Scottish Index of Multiple Deprivation 2020. 2020 [14/05/2025]. Available from: https://www.gov.scot/collections/scottish-index-of-multiple-deprivation-2020/.

29. Ritchie J, Spencer L. Qualitative Data Analysis for Applied Policy Research. In: Bryman A, Burgess R, editors. Analyzing Qualitative Data. London: Routledge; 1994. p. 173–94.

30. UK Parliament. Tobacco and Vapes Bill, HL Bill 89, (2025).

31. Graham-DeMello A, Hoek J, Drew J. How do underage youth access e-cigarettes in settings with minimum age sales restriction laws? A scoping review. BMC Public Health. 2023;23(1):1809.

32. Thomas RE, McLellan J, Perera R. Effectiveness of school-based smoking prevention curricula: systematic review and meta-analysis. BMJ Open. 2015;5(3):e006976.

33. Newton NC, Stapinski LA, Slade T, Sunderland M, Barrett EL, Champion KE, et al. The 7-Year Effectiveness of School-Based Alcohol Use Prevention From Adolescence to Early Adulthood: A Randomized Controlled Trial of Universal, Selective, and Combined Interventions. J Am Acad Child Adolesc Psychiatry. 2022;61(4):520–32.

34. Pulimeno M, Piscitelli P, Colazzo S, Colao A, Miani A. School as ideal setting to promote health and wellbeing among young people. Health Promot Perspect. 2020;10(4):316–24.

35. Jepson R, McAteer J, Williams AJ, Doi L, Buleo A. Developing Public Health Interventions: A Step-by-Step Guide: Sage Publications; 2022.

